# Availability of personal protective equipment and satisfaction of healthcare professionals during COVID-19 pandemic in Ethiopia

**DOI:** 10.1101/2020.10.30.20223149

**Authors:** Wakgari Deressa, Alemayehu Worku, Workeabeba Abebe, Muluken Gizaw, Wondwosson Amogne

## Abstract

Healthcare professionals (HCPs) are at the frontline in the fight against COVID-19 and are at an increased risk of becoming infected with coronavirus. Risk of infection can be minimized by use of proper personal protective equipment (PPE). This study assessed the availability of PPE and satisfaction of HCPs in six public hospitals in Addis Ababa, Ethiopia. A cross-sectional study was conducted from 9^th^ to 26^th^ June 2020. The study hospitals included: Tikur Anbessa Specialized Hospital, Zewditu Memorial Hospital, Ghandi Memorial Hospital, Menelik II Hospital, Yekatit 12 Hospital Medical College and St. Paul Hospital Millennium Medical College. Data were collected using a self-administered questionnaire. Descriptive statistics were used to describe the data and Chi-square test was used to assess the association between the groups. Bivariate and multivariable logistic regression models were used to assess factors associated with the satisfaction level of healthcare workers with regard to the availability and use of proper PPE during the current COVID-19 pandemic. A total of 1,134 (92.3%) valid questionnaires from a possible 1,228 were included in the analysis. The mean (±SD) age of the participants was 30.26±6.43 year and 52.6% were females. Nurses constituted about 40% of the overall sample, followed by physicians (22.2%), interns (10.8%), midwives (10.3%) and others (16.7%). An overall shortage of PPE was reported in all study hospitals. The majority (77%) of the healthcare professionals reported that their hospital did not have adequate PPE. A critical shortage of N95 respirator was particularly reported, the self-reported availability of N95 increased from 13% to 24% before and during COVID-19, respectively. The self-reported use of N95 increased from 9% to 21% before and during COVID-19, respectively. Almost 72% of the respondents were dissatisfied with the availability of PPE in their hospital. The independent predictors of the respondents’ satisfaction level about PPE were male gender (adjusted OR=1.39, 95% CI:1.05-1.85), healthcare workers who reported that PPE was adequately available in the hospital (adjusted OR=7.53, 95% CI:5.08-11.16), and preparedness to provide care to COVID-19 cases (adjusted OR=1.65, 95% CI:1.22-2.12). A critical shortage of appropriate PPE both before and during COVID-19 was identified. The high level of dissatisfaction with the availability of PPE might potentially lead to a lower level of preparedness and readiness to fight against COVID-19. Therefore, urgent efforts are needed to adequately supply the healthcare facilities with appropriate PPE to alleviate the challenges.

## Introduction

The outbreak of coronavirus disease 2019 (COVID-19), caused by the severe acute respiratory syndrome coronavirus 2 (SARS-CoV-2), has been declared as a pandemic by the World Health Organization (WHO) on the 11^th^ of March 2020 [1]. Worldwide, the pandemic has caused over 41 million confirmed cases and more than 1.1 million as of 22^nd^ Oct 2020 2020 [2]. The African continent has the lowest number of globally confirmed cases, standing roughly at 1,685,589 and registering 40,690 deaths. As of 22^nd^ October 2020, Ethiopia has confirmed 91,693 COVID-19 cases, 1,396 deaths, and 45,260 recoveries from over 1,423,505 tests performed to date. At the moment, Ethiopia stood at 4^th^ from Africa in terms of the reported number of confirmed COVID-19 cases next to South Africa (1^st^), Egypt (2^nd^) and Morocco (3^rd^). Thus far the case fatality rate of Ethiopia, which represents 1.5% of the cumulative confirmed COVID-19 cases, is less than the average for Africa (2.4%) and the world (3.3%). Nonetheless, recent reports from the country suggest a spiking rate of coronavirus transmission in the community [3].

Healthcare professionals (HCPs) are at the frontline of defense in combating COVID-19 and they play a critical role, not only in the management of COVID-19 patients, but also in ensuring adequate infection prevention and control (IPC) measures in healthcare settings. As a result, they are at a substantially increased risk of becoming infected with the virus and could potentially contribute to the transmission [4-6]. In Ethiopia, over 1,311 health workers have contracted coronavirus as of 17^th^ September 2020. About 11% of HCPs retrospectively studied in Spain had tested for COVID-19 [7]. Early evidence from countries with the highest mortality rates indicates that healthcare workers are considerably at greater risk of being infected with COVID-19 ranging from 15% to 20% of the infected population and are therefore at a disproportionate risk to the rest of the population [8,9]. For instance, the Italian Regional Reference Laboratories reported that healthcare workers accounted for 10% of 162,000 cases of COVID-19 in the country [10]. Similarly, the US Centers for Disease Control and Prevention reported that accounted for about 11% of all confirmed COVID-19 cases in the United States between [11].

Infection prevention and control (IPC) measures such as the use of appropriate PPE, proper handwashing, and hand hygiene are critical in preventing the transmission and risk of infection of COVID-19 in healthcare settings. The use of appropriate PPE by healthcare workers in particular during the current COVID-19 pandemic is highly recommended and the national and international safety protocols for healthcare workers should be strictly followed [1,12]. Since the initial outbreak report of COVID-19 in China in December 2019, there has been an increasing demand for PPE globally. In many healthcare settings particularly in Africa HCPs have limited access to appropriate PPE to protect their health in many healthcare settings [6]. As a result, many healthcare workers remain concerned about the risk of infection from the SARS-COV-2 due to the shortage of appropriate PPE recommended by WHO, and they have become ill-equipped to care for patients with COVID-19 or other causes, due to acute shortage of appropriate PPE [13].

A lack of PPE puts both HCPs and patients at risk of contracting coronavirus infection. It also presents many HCPs with challenging decisions about whether to care and provide treatment for COVID-19 patients in the absence of effective PPE. In addition, compliance with guidance on the correct use of PPE in healthcare setting is another challenge. On the other hand, the number of COVID-19 cases is rising and the shortages in PPE remains a major concern. The purpose of this study was to assess the self-reported availability and use of PPE as well as satisfaction level of HCPs practicing in public hospitals in Addis Ababa during the current COVID-19 pandemic.

## Methods

### Study area and setting

A hospital-based cross-sectional study was conducted from 9^th^ to 26^th^ June 2020 at six public hospitals in Addis Ababa city administration, three months after the first confirmed COVID-19 case in Ethiopia in March 2020. Addis Ababa city is the most populated urban city in Ethiopia, and is home to about 17% of the urban dwellers in the country. In 2019, the city had a projected population of about 3.6 million and accounted for 3.7% of the total population [14]. The city has the highest number of health infrastructure and medical personnel compared with any city or region in the country. There were 12 hospitals and close to 100 health centers belonging to the public center, and about 25 private hospitals in Addis Ababa city. There were also over 17,000 HCPs in the city, including 2,441 (14%) physicians and 8,172 (47%) nurses by the end of July 2019. Addis Ababa city has the highest rate of COVID-19 cases and deaths in Ethiopia. As of 22^nd^ Oct 2020, a total of 46,570 confirmed COVID-19 confirmed cases and 122 deaths were reported in Addis Ababa [15]. The hospitals selected for the current study were the leading hospitals in the country and provided outpatient and inpatient services for the city residents and patients coming from different parts of the country.

### Study population and sampling

Of the 12 government hospitals in Addis Ababa city administration, the following six were purposively selected based on the relatively higher number of health work forces: Tikur Anbessa Specialized Hospital (TASH), Zewditu Memorial Hospital (ZMH), Ghandi Memorial Hospital (GMH), Menelik II Hospital (MH), Yekatit 12 Hospital Medical College (Y12HMC) and St. Paul Hospital Millennium Medical College (SPHMMC). The study population included all categories of HCPs practicing in the selected hospitals at the time of the survey. In this study, HCP is defined as a healthcare provider in the selected hospital involved in the provision of healthcare services including intern doctors, resident doctors, general practitioners, medical specialists and sub-specialists, health officers, anesthetists, nurses, midwives, laboratory technologists, radiologists, physiotherapists, X-ray and laboratory technicians. The study targeted the HCPs since they are the majority involved in a number of healthcare activities which render them at risk of acquiring and transmitting infections.

Sample size was calculated using a single cross-sectional study design formula based on a 50% prevalence estimate of the availability of PPE in the hospital at 95% confidence level, 4% precision, a design effect of 1.5 and 25% non-response rate. Accordingly, the minimum total sample size targeted for this survey was 1,200 respondents. A mix of purposive and random sampling was applied to select participants based on their availability and willingness to participate in the study. In each hospital, the types and number of wards were initially identified and the number of healthcare workers within each ward was obtained from the human resource department. The sample size allocated to the hospital was distributed to the wards proportional to the size of their healthcare workers. Since it was difficult to obtain the complete list of healthcare workers in each ward at the time of the study, proper random sampling was not followed to select the study participants. Some healthcare workers in particular physicians or nurses were on duty, some were working in different departments in the same hospital or another hospital, and others were reluctant to accept the invitation to participate in the study. The list of the available voluntary healthcare workers was obtained and a simple random sampling was applied to select potential respondents based on the sample size allocated to each ward. All eligible participants who consented to participate were recruited into the study.

### Data collection

Date were collected using structured paper-based self-administered questionnaires that composed of sections on demographic and occupational characteristics of the respondents (e.g., gender, age, education and years of work experience), working unit, availability and practices regarding compliance with usage of PPE (gloves, gowns, facemask, N95 respirator, goggles, face shields, and hair covers), as well as their main concerns and worries about the availability and use of proper PPE during the current COVID-19 pandemic. The satisfaction level of HCPs regarding the availability and use of PPE included four items: (1) I am satisfied with the current availability of PPE in my hospital during the COVID-19, (2) I am satisfied with the current use of PPE by health professionals in my hospital, (3) I am satisfied that the correct PPE (as recommended by WHO) is always available to me when managing suspected or confirmed COVID-19 patient in my hospital, and (4) I am satisfied that the correct PPE (as recommended by WHO) is always available to me when treating non-COVID-19 patients in my hospital. These items were measured using a five-point Likert scale, where 1-strongly dissatisfied, 2-dissatisfied, 3-average, 4-satisfied, and 5-strongly satisfied.

The questionnaire was developed in English based on related literature and available national and international PPE guidelines. A total of 12 experienced data collectors with health backgrounds were involved in data collection. One data collector per hospital was independently recruited and trained for this purpose, while one assistant healthcare workers was recruited from each hospital to facilitate and assist the data collection process. A guideline was developed by the research team to guide the data collectors, assistant healthcare workers and supervisors for data collection, quality assurance of data and ethical conduct during implementation of the survey. The components of the guidelines included sections on selection of respondents, data collection procedures using self-administered questionnaire, and ethical issues including COVID-19 infection prevention measures. Training and orientation on the survey including how to administer the questionnaire were conducted for the data collectors using webinar.

Before handing out the questionnaires to the potential study participants in the selected hospitals, the data collectors introduced themselves to the respondents, build a rapport with them and explained the aims of the study and data collection procedures. After obtaining consent from the participants, the questionnaires were handed out to the respondents and appointed for return to recollect the completed questionnaires. The questionnaires were distributed with a cover letter (consent form), introducing the study and explaining the purpose of the survey, instructions on how to complete the questionnaire, and researchers contact information for any questions the respondent might have. Participants completed the questionnaires by themselves in English language. Data collection took place concurrently in all hospitals. Upon return of the questionnaires, the data collectors checked for completeness and consistency, and incomplete questionnaires were taken back to the respondents for completion as much as possible.

### Statistical analyses

Before data entry, each questionnaire was checked for completeness. Data were entered into the Census Surveys Professional (CSPro) Version 7.2 statistical software package and subsequently exported to SPSS version 23.0 (SPSS Inc., IBM, USA) for cleaning and data analysis. Continuous data were summarized using means and standard deviations, while categorical data were presented as frequency counts and percentages. Descriptive statistics were used to describe the study variables. The Chi-square test was used to assess the association between the groups.

The overall satisfaction score regarding the availability and use of PPE for each respondent was calculated by taking the sum of the scores of the four questions. Responses to these questions were summed to form a total satisfaction score ranging from 4 to 20, with higher scores indicating higher level of satisfaction. Using the total satisfaction score, individuals were classified into two groups: dissatisfied (≤ median score) and satisfied (>median score). The reliability of the questionnaire was measured by Cronbach’s alpha coefficient, and the Cronbach’s alpha for the satisfaction level was 0.769. A bivariate and multivariable binary logistic regression were performed to identify the main factors associated with healthcare professional’s satisfaction level regarding availability and use of PPE. Individuals were classified into two based on their satisfaction level: satisfied group (1), and the rest were placed in the dissatisfied group (0). Odds ratios (ORs) and their 95% confidence intervals (CIs) were used to quantify the associations between potential predictors and outcome variable, satisfaction level. A value of *P*<0.05 was used for all tests of statistical significance.

### Ethical considerations

Ethical clearance for the study was obtained from the Institutional Review Board of the College of Health Sciences at Addis Ababa University (AAU). Permission to undertake this study was obtained from every relevant authority at all levels. Official letters from AAU were written to each hospital to cooperate and participate in the survey. All participants gave their informed consent prior to data collection. Anonymity and data confidentiality were ensured, and no identifiable data from individual participants were collected. All personnel involved in the survey received orientation on COVID-19 infection prevention and control measures.

## Results

### Sample characteristics

From a total of 1,228 questionnaires distributed in six hospitals, 1,146 were completed and returned. Of these, 12 questionnaires were discarded due to missing data, resulting in 1,134 (92%) valid questionnaires for analysis. About 53% were females. Among the respondents reporting age, the mean (±SD) age was 30.3 ±6.4 years, about 58% aged between 20-29 years, and 32% aged 30-39 years (Table 1). The largest number of respondents were from TASH (25%, n=283) and SPHMMC (20.5%, n=233), followed by ZMH (15.6%, n=177) and MH (15.3%, n=174). Nurses constituted about 40% of the overall study participants, followed by physicians (22.2%), interns (10.8%) and midwives (10.3%). Among 252 physicians (22.2%) participated in the study, GPs and resident doctors accounted for 44.8% and 42.9%, respectively, while specialists and sub-specialists consisted the remaining 12.3%. With Gyn&Ob department constituting 17.2% of the respondents, surgical (13.9%), pediatrics (13.1%), medical (13.0%) and OPD (10.5%) departments represented a fairly similar number of study participants. Among the study participants reporting work experience, about 49% and 25% of the respondents reported that they served in the hospital less than 5 years and 5-9 years, respectively. However, the majority of the respondents (67%) at SPHMMC only served less than five years as compared to 28.3% of their counterparts at TASH.

**Table 1.** Characteristics of study participants by hospital

| Characteristics | Hospital, n (%)<sup>*</sup> |  |  |  |  |  | Total, n (%) |
| --- | --- | --- | --- | --- | --- | --- | --- |
|  | *TASH* | *ZMH* | *GMH* | *Y12HMC* | *MH* | *SPHMMC* |  |
| Gender (n=1134) |  |  |  |  |  |  |  |
| Male | 140 (49.5) | 72 (40.7) | 42 (36.5) | 67 (44.1) | 90 (51.7) | 126 (54.1) | 537 (47.4) |
| Female | 143 (50.5) | 105 (59.3) | 73 (63.5) | 85 (55.9) | 84 (48.3) | 107 (45.9) | 597 (52.6) |
| Age group (years)<br>(n=982) |  |  |  |  |  |  |  |
| 20-29 | 83 (42.8) | 98 (60.9) | 55 (51.4) | 69 (46.6) | 98 (62.0) | 166 (77.6) | 569 (57.9) |
| 30-39 | 85 (43.8) | 46 (28.6) | 33 (30.8) | 60 (40.5) | 48 (30.4) | 45 (21.0) | 317 (32.3) |
| ≥40 | 26 (13.4) | 17 (10.6) | 19 (17.8) | 19 (12.8) | 128 (7.6) | 3 (1.4) | 96 (9.6) |
| Mean (±SD) | 32.6 (±7.1) | 30.1 (±6.8) | 31.8 (±7.3) | 31.7 (±6.0) | 29.5 (±5.8) | 27.0 (±4.0) | 30.3 (±6.4) |
| Professional category<br>(n=1134) |  |  |  |  |  |  |  |
| Physician | 79 (27.9) | 39 (22.0) | 17 (14.8) | 35 (23.0) | 39 (22.4) | 43 (18.5) | 252 (22.2) |
| Intern | 17 (6.0) | 36 (20.3) | 7 (6.1) | 12 (7.9) | 29 (16.7) | 22 (9.4) | 123 (10.8) |
| Nurse | 128 (45.8) | 54 (30.5) | 51 (44.3) | 48 (31.6) | 68 (39.1) | 104 (44.6) | 453 (39.9) |
| Midwife | 19 (6.7) | 15 (8.5) | 21 (18.3) | 15 (9.9) | 20 (11.5) | 27 (11.5) | 117 (10.3) |
| Other** | 40 (14.1) | 33 (18.6) | 19 (16.5) | 42 (27.6) | 18 (10.3) | 37 (15.9) | 189 (16.7) |
| Department/Unit<br>(n=1134) |  |  |  |  |  |  |  |
| Gyn&Ob | 20 (7.1) | 35 (19.8) | 56 (48.7) | 22 (14.5) | 33 (19.0) | 29 (12.4) | 195 (17.2) |
| Surgical | 50 (17.7) | 16 (9.0) | 0.0 | 14 (9.2) | 28 (16.1) | 49 (21.0) | 157 (13.8) |
| Pediatrics | 25 (8.9) | 24 (13.6) | 5 (4.3) | 35 (23.0) | 25 (14.4) | 37 (15.9) | 151 (13.3) |
| Medical | 44 (15.6) | 31 (17.5) | 0.0 | 27 (17.8) | 28 (16.1) | 17 (7.3) | 147 (13.0) |
| OPD/Screening/Triage | 41 (14.5) | 17 (9.6) | 16 (13.9) | 21 (13.8) | 19 (10.9) | 30 (9.0) | 144 (12.7) |
| Emergency | 35 (12.4) | 9 (5.1) | 10 (8.7) | 10 (6.6) | 10 (5.7) | 21 (9.0) | 95 (8.4) |
| Anesthesia/OR/IC | 39 (13.8) | 13 (7.3) | 14 (12.2) | 5 (3.3) | 6 (3.4) | 16 (6.9) | 93 (8.2) |
| Other*** | 28 (9.9) | 32 (18.1) | 14 (12.2) | 18 (11.8) | 25 (14.4) | 34 (14.6) | 151 (13.3) |
| Work experience (n=938) |  |  |  |  |  |  |  |
| <5 | 80 (42.1) | 88 (57.1) | 49 (45.0) | 76 (59.4) | 103 (68.2) | 156 (75.7) | 552 (58.8) |
| 5-9 | 81 (42.6) | 39 (25.3) | 40 (36.7) | 42 (32.8) | 32 (21.2) | 44 (21.4) | 278 (29.6) |
| 10-14 | 16 (8.4) | 17 (11.0) | 8 (7.3) | 5 (3.9) | 11 (7.3) | 4 (1.9) | 61 (6.5) |
| 15-34 | 13 (6.8) | 10 (6.5) | 12 (11.0) | 5 (3.9) | 5 (3.3) | 2 (1.0) | 47 (5.0) |
| Total, n (%) | 283 (25.0) | 177 (15.6) | 115 (10.1) | 152 (13.4) | 174 (15.3) | 233 (20.5) | 1134 (100) |

\*TASH: Tikur Anbessa Specialized Hospital; ZMH: Zewditu Memorial Hospital; GMH: Ghandi Memorial Hospital; Y12HMC: Yekatit 12 Hospital
Medical College; MH: Menelik II Hospital; SPHMMC: St. Paul Hospital Millennium Medical College
\*\*Other: Includes anesthetist, pharmacist, health officer, lab technologist and radiographer.
\*\*\*Other: Includes Isolation room/ward, Pharmacy, Oncology, etc.

### Availability of PPE before and during COVID-19

The HCPs were asked the types of PPE that were frequently available in the hospital before and after the COVID-19 pandemic. Table 2 shows the self-reported availability of different PPE by HCPs before and during the COVID-19. Gloves and gowns were reported as the most frequently available PPE in the routine care of patients before and during the pandemic. During the COVID-19 pandemic, the frequent availability of most PPE as reported by the study participants has improved, for example, the frequent availability of surgical facemask and N95 respirator has increased from 59.3% and 12.6% before the pandemic to 82.6% and 24.2% after the pandemic, respectively. The self-reported availability of gloves before and during COVID-19 was >90%, and statistically not significant for any of the hospitals. The availability of gowns for all study hospitals was >60% before and during the COVID-19, with no significant increase during the COVID-19.

**Table 2.** Self-reported frequently available PPEs by hospital before and during COVID-19 (n=1134)

| Frequently available PPE by hospital before and during COVID-19 | Frequently available PPE, % |  |  |  |  |  |
| --- | --- | --- | --- | --- | --- | --- |
|  | *Glove* | *Gown* | *Facemask* | *N95 respirator* | *Eye protection* | *Hair cover* |
| TASH (n=283)* |  |  |  |  |  |  |
| Before | 92.9 | 73.5 | 77.4 | 23.3 | 21.6 | 19.1 |
| During | 93.3 | 70.3 | 86.9 | 30.0 | 27.2 | 20.8 |
| *P-value* | 0.868 | 0.400 | <0.001 | 0.071 | 0.117 | 0.559 |
| ZMH (n=177) |  |  |  |  |  |  |
| Before | 96.6 | 65.5 | 59.3 | 5.1 | 14.7 | 11.3 |
| During | 95.5 | 67.2 | 83.1 | 16.9 | 22.0 | 19.8 |
| *P-value* | 0.586 | 0.736 | <0.001 | <0.001 | 0.074 | 0.028 |
| GMH (n=115) |  |  |  |  |  |  |
| Before | 97.4 | 76.5 | 67.8 | 3.5 | 25.2 | 21.7 |
| During | 96.5 | 76.5 | 87.8 | 13.0 | 33.9 | 19.1 |
| *P-value* | 0.701 | 1.00 | <0.001 | 0.008 | 0.149 | 0.624 |
| YHMC (n=152) |  |  |  |  |  |  |
| Before | 92.8 | 84.9 | 42.1 | 12.5 | 11.8 | 13.2 |
| During | 92.1 | 82.9 | 87.5 | 28.9 | 15.1 | 13.8 |
| *P-value* | 0.828 | 0.640 | <0.001 | <0.001 | 0.401 | 0.867 |
| MH (n=174) |  |  |  |  |  |  |
| Before | 93.1 | 73.0 | 48.3 | 5.7 | 8.6 | 6.9 |
| During | 90.8 | 67.8 | 76.4 | 9.2 | 12.1 | 6.9 |
| *P-value* | 0.431 | 0.291 | <0.001 | 0.221 | 0.291 | 1.00 |
| SPHMMC (n=233) |  |  |  |  |  |  |
| Before | 92.3 | 63.5 | 52.3 | 15.0 | 26.6 | 21.9 |
| During | 87.1 | 61.8 | 76.0 | 36.1 | 46.4 | 23.2 |
| *P-value* | 0.067 | 0.702 | <0.001 | <0.001 | <0.001 | 0.721 |
| All hospitals (n=1134) |  |  |  |  |  |  |
| Before | 93.8 | 72.0 | 59.3 | 12.6 | 18.6 | 16.0 |
| During | 92.2 | 70.0 | 82.6 | 24.2 | 27.1 | 17.9 |
| *P-value* | 0.118 | 0.309 | <0.001 | <0.001 | <0.001 | 0.240 |

\* TASH: Tikur Anbessa Specialized Hospital; ZMH: Zewditu Memorial Hospital; GMH: Ghandi Memorial Hospital; YHMC: Yekatit 12 Hospital Medical College; MH: Menelik II Hospital; SPHMMC: St. Paul Hospital Millennium Medical College.

The availability of facemask and N95 respirator showed a statistically significant increase during COVID-19 as compared to the pre-COVID-19 (*P*<0.001). Similarly, the use of eye protection (goggles and face shield) has increased from 18.6% before the pandemic to 27.1% during the pandemic (*P*<0.001), but only a steady increase was observed in the availability of hair covers during the pandemic as compared with the time before COVID-19 (*P*=0.240). This study found a major variation among the study hospitals with regard to the frequently available PPE before and after COVID-19. The frequent availability of N95 respirator during the pandemic was reported by 36% of the participants from SPHMMC and 30% from TASH as compared with 9.2% at MH and 13% at GMH. Even simple hand sanitizer was in short supply in some hospitals as reported by some respondents.

### Use of PPE before and during COVID-19

The HCPs were asked the types of PPE that were frequently used in the hospital before and after the COVID-19 pandemic. Table 3 presents the self-reported frequently used PPE by HCPss before and during COVID-19. Gloves and gowns were identified as the most frequently used PPE in the hospital before and during the COVID-19 pandemic. The use of gloves by all HCPs was above 90%, while the self-reported use by other healthcare workers before and after COVID-19 was relatively lower than others, despite showing some improvement during COVID-19. Likewise, the self-reported use of gowns remained not statistically significant before and during COVID-19, whereas its use rate remained less than 80% for the different categories of HCPs. The use of surgical facemask has increased from 47.2% before the pandemic to 85.7% during the pandemic for all HCPs (*P*<0.001).

**Table 3.** Self-reported frequently used PPE by healthcare professionals before and during COVID-19 (n=1134)

| PPE use by professional category<br>before and during COVID-19 | Frequently used PPE, % |  |  |  |  |  |
| --- | --- | --- | --- | --- | --- | --- |
|  | *Glove* | *Gown* | *Facemask* | *N95 respirator* | *Eye protection* | *Hair cover* |
| Use by physician (n=252) |  |  |  |  |  |  |
| Before | 94.0 | 66.3 | 48.4 | 15.9 | 17.8 | 11.9 |
| During | 94.8 | 69.0 | 89.7 | 23.0 | 17.1 | 14.7 |
| *P-value* | 0.697 | 0.505 | <0.001 | 0.043 | 0.815 | 0.358 |
| Use by intern (n=123) |  |  |  |  |  |  |
| Before | 91.1 | 62.6 | 36.6 | 3.3 | 4.1 | 4.1 |
| During | 92.7 | 62.6 | 92.7 | 13.0 | 14.6 | 2.4 |
| *P-value* | 0.641 | 1.00 | <0.001 | 0.005 | 0.004 | 0.472 |
| Use by nurse (n=453) |  |  |  |  |  |  |
| Before | 92.3 | 74.0 | 55.0 | 9.1 | 12.1 | 17.9 |
| During | 91.8 | 71.5 | 82.1 | 22.3 | 24.3 | 22.3 |
| *P-value* | 0.806 | 0.412 | <0.001 | <0.001 | <0.001 | 0.097 |
| Use by midwife (n=117) |  |  |  |  |  |  |
| Before | 95.7 | 67.5 | 41.0 | 5.1 | 16.2 | 22.2 |
| During | 94.0 | 68.4 | 79.5 | 21.4 | 40.2 | 29.9 |
| *P-value* | 0.553 | 0.889 | <0.001 | <0.001 | <0.001 | 0.180 |
| Use by others (n=189)* |  |  |  |  |  |  |
| Before | 81.5 | 79.4 | 37.6 | 6.3 | 4.2 | 7.9 |
| During | 86.2 | 77.8 | 88.4 | 21.2 | 19.0 | 14.3 |
| *P-value* | 0.208 | 0.707 | <0.001 | <0.001 | <0.001 | 0.049 |
| Use by all categories (n=1134) |  |  |  |  |  |  |
| Before | 91.1 | 71.3 | 47.2 | 9.1 | 11.6 | 13.8 |
| During | 91.9 | 70.7 | 85.7 | 21.2 | 22.4 | 17.9 |
| *P-value* | 0.498 | 0.781 | <0.001 | <0.001 | <0.001 | 0.008 |

\*Other: Includes anesthetist, pharmacist, health officer, lab technologist and radiographer.

The use of N95 respirator has also increased from 9.1% before the pandemic to 21.2% after the pandemic (*P*<0.001). Similarly, the use of eye protection (goggles and face shield) has increased from 11.6% before the pandemic to 22.4% during the pandemic (*P*<0.001). A statistically significant increase in the percentage of respondents reporting the frequent use of hair covers during the pandemic as compared with the time before COVID-19 was also reported (*P*=0.008). The self-reported use of N95 respirator was the highest for physicians than other even before (16%) and after (23%) COVID-19, while the least use of N95 respirator was reported by interns. Overall, the self-reported use of N95 respirator was lower than other PPE except the use of hair cover. Although there was an increase in the self-reported use of hair cover during COVID-19, its use was generally very low and the difference was not statistically significant regarding its use by the different categories of HCPs.

With regard to the types of PPE used during their last interaction with a patient, the majority of the HCPs reported the use of gloves (91.2%), gowns (72.4%), and facemasks (86.8%), with about 24%, 19% and 18% reporting N95 respirator, eye protection and hair dresses, respectively (Table 4).

**Table 4.** Self-reported use of PPE during last interaction with a patient by healthcare professionals (n=1080)

| Characteristics | Last used PPE, % |  |  |  |  |  |
| --- | --- | --- | --- | --- | --- | --- |
|  | Glove | Gown | Facemask | N95 respirator | Eye protection | Hair cover |
| Gender |  |  |  |  |  |  |
| Male | 90.9 | 72.2 | 83.7 | 27.0 | 18.3 | 15.0 |
| Female | 88.5 | 70.1 | 86.7 | 20.1 | 18.7 | 19.4 |
| *P-value* | 0.208 | 0.461 | 0.152 | 0.007 | 0.853 | 0.053 |
| Professional category |  |  |  |  |  |  |
| Physician | 93.4 | 68.4 | 89.3 | 24.2 | 14.3 | 12.7 |
| Intern | 94.7 | 65.8 | 94.7 | 21.9 | 15.8 | 5.3 |
| Nurse | 89.5 | 73.1 | 81.3 | 22.4 | 20.1 | 22.4 |
| Midwife | 93.7 | 71.2 | 84.7 | 25.2 | 34.2 | 28.8 |
| Other* | 79.2 | 73.2 | 83.6 | 24.6 | 12.6 | 12.0 |
| *P-value* | <0.001 | 0.453 | 0.002 | 0.939 | <0.001 | <0.001 |
| Department/Unit |  |  |  |  |  |  |
| Gyn&Ob | 96.3 | 70.6 | 87.8 | 19.3 | 24.6 | 20.3 |
| Surgical | 96.0 | 71.3 | 84.0 | 32.0 | 24.0 | 27.3 |
| Pediatrics | 87.3 | 69.0 | 86.6 | 23.2 | 16.2 | 9.2 |
| Medical | 89.9 | 75.5 | 89.9 | 19.4 | 14.4 | 10.8 |
| OPD/Screening/Triage | 79.6 | 75.2 | 80.3 | 22.6 | 15.3 | 11.7 |
| Emergency | 93.4 | 81.3 | 91.2 | 20.9 | 17.6 | 14.3 |
| Anesthesia/OR/IC | 94.3 | 59.2 | 86.4 | 30.7 | 22.7 | 38.6 |
| Other** | 80.8 | 66.4 | 78.1 | 21.9 | 12.3 | 11.6 |
| *P-value* | <0.001 | 0.034 | 0.036 | 0.090 | 0.032 | <0.001 |
| Hospital*** |  |  |  |  |  |  |
| TASH | 94.7 | 74.2 | 87.5 | 24.2 | 13.6 | 22.0 |
| ZMH | 85.5 | 62.8 | 84.9 | 14.0 | 12.2 | 13.4 |
| GMH | 93.0 | 79.1 | 91.3 | 8.7 | 20.0 | 16.5 |
| Y12HMC | 84.5 | 81.1 | 90.5 | 28.4 | 12.8 | 13.5 |
| MH | 89.4 | 72.0 | 77.0 | 14.3 | 11.8 | 6.8 |
| SPHMMC | 88.6 | 62.3 | 82.3 | 40.9 | 37.3 | 25.5 |
| *P-value* | 0.006 | <0.001 | 0.003 | <0.001 | <0.001 | <0.001 |
| Overall use of PPE | 89.6 | 71.1 | 85.3 | 23.4 | 18.5 | 17.3 |

\*Other: Includes anesthetist, pharmacist, health officer, lab technologist and radiographer.
\*\*Other: Includes Isolation room/ward, Pharmacy, Oncology, etc.
\*\*\* TASH: Tikur Anbessa Specialized Hospital; ZMH: Zewditu Memorial Hospital; GMH: Ghandi Memorial Hospital; YHMC: Yekatit 12 Hospital Medical College; MH: Menelik II Hospital; SPHMMC: St. Paul Hospital Millennium Medical College.

### Self-reported satisfaction level of healthcare professionals about PPE

Table 5 shows the satisfaction level of HCPs with regard to the current availability and use of PPE in the study hospitals, and 54.7% (n=584) and 17.5% (n=187) of the respondents reported that they were unsatisfied or somewhat unsatisfied with the availability of PPE, respectively. Similarly, 48.8% (n=521) and 20% (n=213) of the participants self-reported that they were unhappy or somewhat unhappy with the current use of PPE by health professionals in the hospital. Overall, only 12% or less of the respondents expressed their opinion that they were satisfied or somewhat satisfied with the current availability or use of PPE at their hospitals.

**Table 5.** Satisfaction level about the availability and use of PPE in the study hospitals by professional category (n=1067)

| Variable | Professional category, n (%) |  |  |  |  | Total, n (%) |
| --- | --- | --- | --- | --- | --- | --- |
|  | *Physician* | *Intern* | *Nurse* | *Midwife* | *Other\** |  |
| I am satisfied with the current availability of PPE in my hospital during the COVID-19 |  |  |  |  |  |  |
| Strongly satisfied | 4 (1.7) | 1 (0.8) | 8 (1.9) | 2 (1.8) | 3 (1.7) | 18 (1.7) |
| Satisfied | 18 (7.5) | 1 (0.8) | 43 (10.2) | 13 (11.9) | 15 (8.5) | 90 (8.4) |
| Average | 40 (16.7) | 26 (22.0) | 62 (14.7) | 22 (20.2) | 38 (21.5) | 188 (17.6) |
| Dissatisfied | 54 (22.5) | 17 (14.4) | 71 (16.8) | 18 (16.5) | 27 (15.3) | 187 (17.5) |
| Strongly dissatisfied | 124 (51.7) | 73 (61.9) | 239 (56.5) | 54 (49.5) | 94 (53.1) | 584 (54.7) |
| I am satisfied with the current use of PPE by health professionals in my hospital |  |  |  |  |  |  |
| Strongly satisfied | 4 (1.7) | 1 (0.8) | 9 (2.1) | 3 (2.8) | 4 (2.3) | 21 (2.0) |
| Satisfied | 27 (11.3) | 6 (5.1) | 42 (9.9) | 18 (16.5) | 17 (9.6) | 110 (10.3) |
| Average | 39 (16.3) | 26 (22.0) | 80 (18.9) | 20 (18.3) | 37 (20.9) | 202 (18.9) |
| Dissatisfied | 77 (32.1) | 18 (15.3) | 71 (16.8) | 17 (15.6) | 30 (16.9) | 213 (20.0) |
| Strongly dissatisfied | 93 (38.8) | 67 (56.8) | 221 (52.2) | 51 (46.8) | 89 (50.3) | 521 (48.8) |
| I am satisfied that the correct PPE (as recommended by WHO) is always available to me when managing suspected or confirmed COVID-19 patient in my hospital |  |  |  |  |  |  |
| Strongly satisfied | 16 (6.7) | 10 (8.5) | 65 (15.4) | 13 (11.9) | 28 (15.8) | 132 (12.4) |
| Satisfied | 73 (30.4) | 16 (13.6) | 83 (19.6) | 22 (20.2) | 40 (22.6) | 234 (21.9) |
| Average | 27 (11.3) | 13 (11.0) | 50 (11.8) | 6 (5.5) | 20 (11.3) | 116 (10.9) |
| Dissatisfied | 54 (22.5) | 27 (22.9) | 137 (32.4) | 34 (31.2) | 42 (23.7) | 294 (27.6) |
| Strongly dissatisfied | 70 (29.2) | 52 (44.1) | 88 (20.8) | 34 (31.2) | 47 (26.6) | 291 (27.3) |
| I am satisfied that the correct PPE (as recommended by WHO) is always available to me when treating non-COVID-19 patients in my hospital |  |  |  |  |  |  |
| Strongly satisfied | 14 (5.8) | 7 (5.9) | 63 (14.9) | 12 (11.0) | 25 (14.1) | 121 (11.3) |
| Satisfied | 79 (32.9) | 20 (16.9) | 110 (26.0) | 28 (25.7) | 44 (24.9) | 281 (26.3) |
| Average | 17 (7.1) | 10 (8.5) | 84 (19.6) | 8 (7.3) | 16 (9.0) | 96 (9.0) |
| Dissatisfied | 67 (27.9) | 32 (27.1) | 112 (26.5) | 33 (30.3) | 46 (26.0) | 290 (27.2) |
| Strongly dissatisfied | 63 (26.3) | 49 (41.5) | 93 (22.0) | 28 (25.7) | 46 (26.0) | 279 (26.1) |
| *Total, n (%)* | *240 (22.5)* | *118 (11.1)* | *423 (39.6)* | *109 (10.2)* | *177 (16.6)* | *1067 (100)* |

\*Other: Includes anesthetist, pharmacist, health officer, lab technologist and radiographer.

About 28% (n=294) and 27% (n=291) of all the respondents self-reported that they were dissatisfied or strongly dissatisfied, respectively, about the availability of the correct PPE in their hospital, as recommended by WHO, for managing suspected/confirmed COVID-19 patients (Table 5). It is only about one-third of the respondents who agreed or strongly agreed about the availability of PPEs in their hospital for managing COVID-19 patients as recommended by WHO.

Generally, more than half of the different healthcare professional categories reported that they disagreed or strongly disagreed about the statement on the availability of correct PPE in the hospital for managing COVID-19 patients as per the WHO recommendation, ranging from about 51% by physicians and 66% by interns. About 54% and 17.5% of the respondents reported that they were unsatisfied or somewhat unsatisfied with the availability of PPE, respectively. The overwhelming majority of interns (76.8%), physicians (72.3%) and nurses (72.9%) were unsatisfied with the current availability of PPE in the study hospitals. Only 10% of the respondents expressed their opinion that they were satisfied or somewhat satisfied with the current availability of PPE at their hospitals.

This study also assessed the level of preparedness of HCPs to provide direct clinical care to COVID-19 patients. Only 5.2% and 32.8% of the participants felt they were completely prepared or somewhat prepared to provide direct clinical care to COVID-19 patients, respectively. Overall, the majority (77%, n=872/1134) of the participants perceived that the PPE currently available to them at their hospital was inadequate to keep them safe from infection when managing suspected or confirmed COVID-19 patients. The mean and the SD of the satisfaction scores of the four items regarding the availability and use of PPE were calculated. Table 6 shows the degree of satisfaction scores of all respondents. The first two items had a score of <2 (1.85±1.13 and 1.97±1.13), indicating strong dissatisfaction of the HCPs, while the remaining two items had a mean score of between 1.5 and 2.0 (2.65±1.40 and 2.70±1.39), showing the dissatisfaction of the study participants. The overall score was 2.29±0.97, showing that the majority of the respondents reported that they were dissatisfied with the availability and use of PPE in the study hospitals.

**Table 6.**
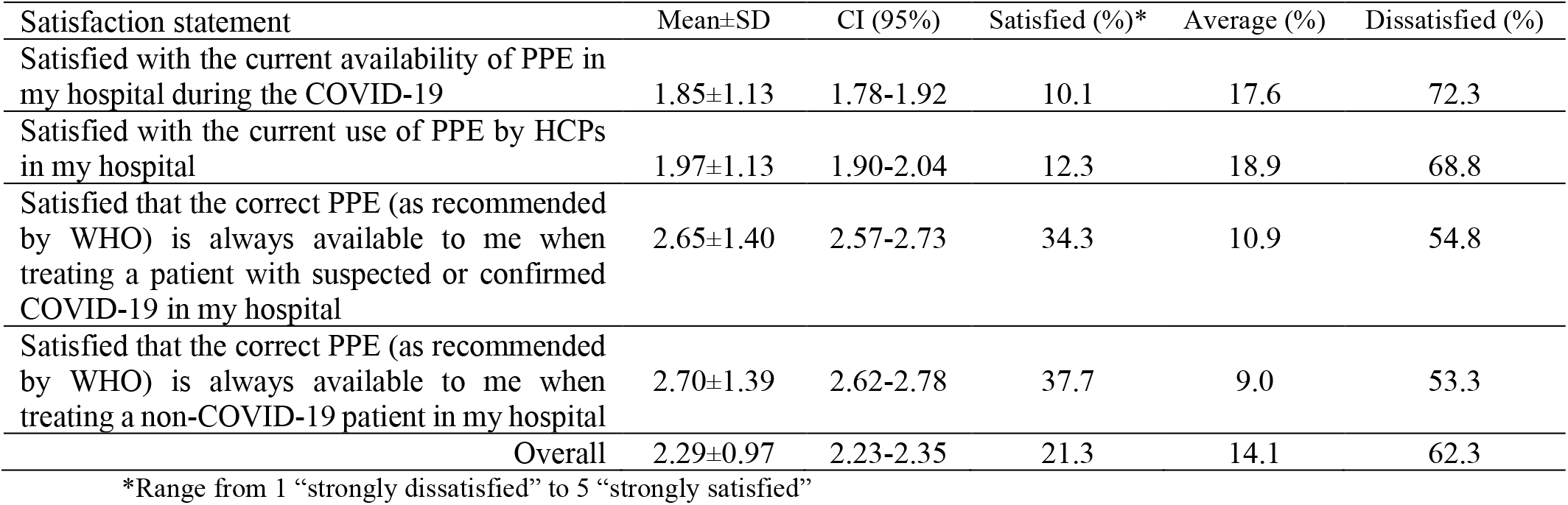
Descriptive statistics of PPE satisfaction of healthcare professionals (n=1067)

### Bivariate and multivariable analyses of PPE self-reported satisfaction

The total satisfaction score regarding the availability and use of PPE for each respondent was used as dependent variable and dichotomized into two groups: dissatisfied (≤median score) and satisfied (>median score). This dependent variable was further subjected to bivariate and multivariable binary logistic regression analyses using eight potential independent variables (gender, medical profession, working unit, hospital, whether received training in PPE during the COVID-19 pandemic, whether used any ‘homemade’ or ‘creative’ PPE during COVID-19, whether they reported that adequate PPE was available to protect them from risk of infection while managing suspected/confirmed COVID-19 patients, and preparedness to tackle COVID-19).

Table 7 shows the relationship between the respondents PPE satisfaction level and independent factors for both bivariate and multivariable logistic regression analyses. In the bivariate logistic regression, the odds of satisfaction with the availability of PPE among males were 1.37 times higher than females (OR=1.37, 95% CI:1.08-1.75, *P*=0.010). However, nurses were less likely to be satisfied with the availability and use of PPE than physicians (OR=0.48, 95% CI:0.29-0.77, *P*=0.003). The HCPs at MH were less likely to be satisfied with the availability and use of PPE in their hospital than those in SPHMMC (OR=0.46, 95% CI:0.30-0.72, *P*=0.011). The odds of satisfaction among those respondents who reported that PPE was adequately available to protect themselves from the risk of infection when managing suspected or confirmed COVID-19 cases were 8.31 times higher than among those who said ‘no’ (OR=8.31, 95% CI:5.84-11.82, *P*<0.001). The odds of satisfaction among those health workers who used any ‘homemade’ or ‘creative’ PPE such as homemade fabric, face covering clothes or sewed cotton masks (OR=2.01, 95% CI:1.53-2.66, *P*<0.001) and those who reported that they were prepared to provide direct care to suspected or confirmed COVID-19 cases (OR=1.45, 95% CI:1.12-1.87, *P*=0.004) were higher than among other healthcare workers.

**Table 7.** Factors associated with satisfaction of healthcare professionals regarding the availability and use of PPE using binary logistic regression analyses (n=1067)

| Predictor | Satisfaction level, n (%) |  | Crude OR (95% CI)* | P-value | Adjusted OR (95% CI) | P-value |
| --- | --- | --- | --- | --- | --- | --- |
|  | Dissatisfied (≤median score) | Satisfied (>median score) |  |  |  |  |
| Gender |  |  |  |  |  |  |
| Male | 247 (48.8) | 259 (51.2) | 1.37 (1.08-1.75) | 0.010 | 1.39 (1.05-1.85) | 0.023 |
| Female | 318 (56.7) | 243 (43.3) | 1.0** |  | 1.0 |  |
| Professional category |  |  |  |  |  |  |
| Physician | 119 (49.6) | 121 (50.4) | 1.0 |  | 1.0 |  |
| Intern | 79 (66.9) | 39 (33.1) | 0.98 (0.67-1.45) | 0.931 | 0.79 (0.47-1.34) | 0.386 |
| Nurse | 221 (52.2) | 202 (47.8) | 0.48 (0.29-0.77) | 0.003 | 1.05 (0.72-1.53) | 0.795 |
| Midwife | 59 (54.1) | 50 (45.9) | 0.88 (0.62-1.26) | 0.489 | 1.37 (0.74-2.56) | 0.321 |
| Other*** | 87 (49.2) | 90 (50.8) | 0.82 (0.51-1.32) | 0.414 | 0.94 (0.58-1.52) | 0.800 |
| Department/Unit |  |  |  |  |  |  |
| Gyn&Ob | 109 (58.9) | 76 (41.1) | 1.0 |  | 1.0 |  |
| Surgical | 81 (54.0) | 69 (46.0) | 0.65 (0.42-1.01) | 0.055 | 0.55 (0.29-1.04) | 0.064 |
| Pediatrics | 76 (54.3) | 64 (45.7) | 0.79 (0.50-1.26) | 0.326 | 0.89 (0.51-1.55) | 0.683 |
| Medical | 67 (50.4) | 66 (49.6) | 0.79 (0.49-1.25) | 0.311 | 0.84 (0.47-1.50) | 0.558 |
| OPD/Screening/Triage | 76 (55.9) | 60 (44.1) | 0.92 (0.57-1.47) | 0.726 | 0.73 (0.41-1.30) | 0.281 |
| Emergency | 39 (43.3) | 51 (56.7) | 0.74 (0.46-1.18) | 0.203 | 0.62 (0.36-1.07) | 0.086 |
| Anesthesia/OR/IC | 47 (53.4) | 41 (46.6) | 1.22 (0.72-2.07) | 0.460 | 0.72 (0.39-1.35) | 0.306 |
| Other*** | 70 (48.3) | 75 (51.7) | 0.81 (0.48-1.38) | 0.448 | 0.80 (0.44-1.48) | 0.484 |
| Hospital**** |  |  |  |  |  |  |
| TASH | 124 (45.8) | 147 (54.2) | 1.55 (1.08-2.22) | 0.017 | 1.08 (0.75-1.34) | 0.750 |
| ZMH | 88 (54.0) | 75 (46.0) | 1.11 (0.74-1.67) | 0.609 | 1.42 (0.83-2.44) | 0.203 |
| GMH | 50 (44.2) | 63 (55.8) | 1.65 (1.04-2.60) | 0.033 | 1.69 (1.05-2.71) | 0.031 |
| Y12HMC | 60 (42.9) | 80 (57.1) | 1.74 (1.13-2.67) | 0.033 | 0.45 (0.28-0.72) | 0.001 |
| MH | 119 (23.9) | 42 (26.1) | 0.46 (0.30-0.72) | 0.011 | 0.83 (0.54-1.25) | 0.366 |
| SPHMMC | 124 (56.6) | 95 (43.9) | 1.0 | 0.001 | 1.0 |  |
| Received training in PPE since COVID-19 pandemic |  |  |  |  |  |  |
| Yes | 212 (50.6) | 207 (49.4) | 0.86 (0.67-1.10) | 0.215 | 1.00 (0.75-1.34) | 0.976 |
| No | 353 (54.5) | 295 (45.5) | 1.0 |  | 1.0 |  |
| Reported that PPE was adequately available to protect the risk of coronavirus infection |  |  |  |  |  |  |
| Yes | 45 (17.6) | 210 (82.4) | 8.31 (5.84-11.82) | <0.001 | 7.53 (5.08-11.16) | <0.001 |
| No | 520 (64.0) | 292 (36.0) | 1.0 |  | 1.0 |  |
| Used any 'homemade', or 'creative' PPE |  |  |  |  |  |  |
| Yes | 116 (40.3) | 172 (59.7) | 2.01 (1.53-2.66) | <0.001 | 1.27 (0.90-1.78) | 0.170 |
| No | 449 (57.6) | 330 (42.4) | 1.0 |  | 1.0 |  |
| Prepared to provide direct care to COVID-19 cases |  |  |  |  |  |  |
| Yes | 167 (46.8) | 190 (53.2) | 1.45 (1.12-1.87) | 0.004 | 1.65 (1.22-2.12) | 0.001 |
| No | 398 (56.1) | 312 (43.9) | 1.0 |  | 1.0 |  |
| Total | 565 (53.0) | 502 (47.0) | 1067 |  |  |  |

In the multivariable logistic regression analysis (Table 7), which was performed using eight independent variables in the model, the odds of satisfaction with availability of PPE among males were again 1.39 times higher than female healthcare workers (OR=1.39, 95% CI:1.05-1.85, *P*=0.023). The odds of satisfaction among HCPs from GMH were 1.69 times higher than among those working at SPHMMC (OR=1.69, 95% CI:1.05-2.71, *P*=0.031), while healthcare workers at MH were less likely to be satisfied with PPE than those at SPHMMC (OR=0.45, 95% CI:0.28-0.72, *P*=0.001). The odds of satisfaction among those healthcare workers who reported that PPE was adequately available to protect themselves from the risk of infection when managing suspected or confirmed COVID-19 cases were 7.53 times higher than among those who reported inadequate PPE (OR=7.53, 95% CI:5.08-11.16, *P*<0.001). The odds of satisfaction among HCPs who reported that they were prepared to provide direct care to suspected or confirmed COVID-19 cases were higher than other healthcare workers (OR=1.65, 95% CI:1.22-2.12, *P*=0.001). The factors such as medical profession, medical unit, training in PPE after COVID-19, and use of any ‘homemade’, or ‘creative’ PPE in the hospital did not have significant influence on the satisfaction level of HCPs regarding the availability and use of PPE. The multivariable binary logistic regression model presented in Table 7 had goodness-of-fit under the Hosmer-Lemeshow test (χ^2^=5.35, *P*=0.720), and the full model containing all predictors was statistically significant χ^2^ (21, N=1067) = 231.56, *P*<0.001. The model as a whole explained between 19.5% (Cox-Snell R^2^) and 26% (Nagelkerke R^2^) of the variance in satisfaction level.

## Discussion

This study aimed to investigate the availability and use of PPE among 1,134 HCPs working in six public hospitals during the early stage of COVID-19 in Addis Ababa, Ethiopia. Our findings showed limited access to appropriate and sufficient PPE to health workers in the care of COVID-19 and non-COVID-19 patients before and after COVID-19. Shortages of appropriate PPE for HCPs irrespective of the hospitals they were serving is observed. This raises a concern regarding the availability and use of proper PPE in the hospitals and the challenges of healthcare workers to combat COVID-19 infection. Despite these concerns, the HCPs continue to work during COVID-19. Though there is a global shortage, HCPs must be equipped with appropriate PPE that they need to practice their clinical role with confidence. Shortage of healthcare workers is already significant amidst the national effort against COVID-19. In the previous studies, inadequate personal protection during the management of suspected or confirmed COVID-19 patients, proximity to patients infected by the virus and prolonged exposure to the infected environment were cited as reasons for the health care workers becoming infected with the virus [16,17].

Lack of appropriate PPE itself can put the HCPs at risk of contracting the virus and infecting other healthcare workers and their family. Although this problem did not only exist in Ethiopia, it was also reported from China [18] and other countries. In one study in Jordan, only 18.5% of frontline doctors reported that all PPE were available and most shortage was reported in protective facemasks [19]. Several studies emphasized that adequate training, proper use and uninterrupted availability of adequate PPE give HCPs a minimal risk of infection when treating suspected or confirmed cases of COVID-19 [18,20,21]. A study in China found that of 420 doctors and nurses deployed to frontline work at Wuhan hospitals between January and April 2020 none of them contracted COVID-19 after receiving training in proper use of PPE and provided with appropriate PPE [22]. A study from Hong Kong demonstrated that correct use of PPE by healthcare workers was associated with no cases of infection over a 42-day observation period [23]. Studies have also revealed that the risk of SARS-CoV-2 infection is significantly higher particularly among frontline HCPs with inadequate PPE caring for suspected or confirmed COVID-19 patients [24].

The shortage of PPE is particularly concerning for the commonly used N95 respirators. However, recommendations are currently available to use surgical or medical masks when N95 is in short supply. A recent systematic review and meta-analysis showed that medical masks are not inferior to N95 respirators for protecting healthcare workers against viral respiratory infections during routine care and non–aerosol-generating procedures, but the researchers strongly recommended preservation of N95 respirators for high-risk, aerosol-generating procedures during COVID-19 when its supply is inadequate [25]. In response to the shortage of appropriate PPE, studies showed that the scarcity could be mitigated through proper re-use or extended use techniques [26,27].

Evidence indicates that N95 respirators maintain their protection when used for extended periods [28] although using them for longer than four hours is not recommended due to increased discomfort [28,29]. The choice of PPE is also dependent on the level of protection provided by PPE and the risk of exposure, thus understanding them is the key in choosing appropriate PPE [30]. In this regard, the WHO IPC recommendations have proven to be an invaluable resource and were quickly adopted and implemented in many countries in preparing their response to the COVID-19 pandemic [31]. As a result, the WHO guidance on the rational use of PPE for COVID-19 has provided appropriate criteria on how to select and use appropriate PPE in different settings when PPE is in short supply [13].

The current study gives a first impression of the satisfaction level of HCPs with regard to the availability and use of proper PPE during the COVID-19 pandemic. The findings show that the HCPs had an overall low level of satisfaction with the availability and use of appropriate PPE in their hospital. The healthcare system, which was already affected by the widespread shortage of HCPs, will be further affected by the dissatisfied health workforce. Currently, there is limited evidence on the satisfaction of healthcare workers about the availability and use of PPE. A recent study conducted in Ethiopia reported that 75% of the healthcare workers in hospitals felt unsafe about their work environment and only <30% reported that they had access to proper PPE in the hospitals [32].

The multivariable logistic regression analysis showed that the satisfaction of HCPs regarding the availability and use of PPE were affected by different factors, such as gender, hospital, perception that PPE is adequately available, and preparedness to provide direct care to suspected or confirmed COVID-19 cases. Male healthcare workers reported statistically significant higher satisfaction level with PPE than female health workers. Among the healthcare workers, those who reported that PPE was adequately available to protect themselves from the risk of infection when managing COVID-19 patients rather than those who reported the inadequacy of PPE in their hospitals had statistically significant level of higher satisfaction about PPE. As there is limited published research on the relationship between healthcare workers satisfaction level with regard to the availability and use of appropriate PPE and associated factors, this study contributes additional knowledge in this area of research.

Finally, this study had some limitations. First, the study might be affected by selection bias. Second, the study focused on more general populations of HCPs similar to other studies [33,34] rather than healthcare workers who might have direct contact with COVID-19 patients [35]. Relying solely on respondents to determine the availability and use of PPE can introduce recall bias. Lastly, the results of this study are based on a self-reported questionnaire using a cross-sectional design, and the self-reported response might not represent actual or genuine answers. Despite these limitations, the results obtained provide important information to guide the efforts to avail appropriate PPE and optimize its use for effectively reducing the risk of COVID-19 infection among HCPs through implementing appropriate IPC measures.

## Conclusions

In conclusion, this study has illuminated the level of the availability and use of PPE by HCPs working at hospitals, and identified a critical shortage of appropriate PPE both before and during COVID-19. The availability of N95 respirator was particularly insufficient, and the use of goggle and gown were unsatisfactory, which might increase the risk of COVID-19 infection among HCPs. The study shows that the HCPs had an overall low level of satisfaction with the current availability and use of PPE in their hospital, which might potentially lead to a lower level of preparedness and readiness among health workers to fight against COVID-19 infection. With the current critical shortages of PPE in hospitals, the ongoing widespread COVID-19 pandemic in Ethiopia could result in devastating consequences. The findings provide considerable insights into the importance of urgent need and concerted efforts to adequately supply the healthcare facilities with appropriate PPE to alleviate the current challenges during the COVID-19 pandemic. Preventing the risks of COVID-19 infection among HCPs through providing proper and adequate PPE should be strengthened and needs to be a top priority for Ministry of Health and the Government of Ethiopia.

## Data Availability

Data will be available upon request

## Acknowledgments

The authors are grateful to the participating hospitals and their healthcare staff for committing their time and voluntarily filling in the questionnaire. They are also thankful to all data collectors and logistics facilitators for their time and commitment.

## Ethical approval and consent to participate

Ethical clearance was obtained from the Institutional Review Board of the College of Health Sciences at Addis Ababa University. All participants gave their informed consent.

## Contributors

Conceptualization: WD, AW, WAA, WA

Designing of the study: WD, AW, MG, WAA, WA

Data curation: WD, AW

Statistical analysis: WD, AW

Field supervision: MG, WD, AW

Writing – original draft: WD, AW

Writing – review & editing: WD, AW, MG, WAA, WA

Substantial contribution to the interpretation of the data: WD, AW, MG, WAA, WA

All the authors read and approved the final manuscript.

